# Adaptive human behavior and delays in information availability autonomously modulate epidemic waves

**DOI:** 10.1101/2024.11.23.24317838

**Authors:** Md Shahriar Mahmud, Solomon Eshun, Baltazar Espinoza, Claus Kadelka

## Abstract

The recurrence of epidemic waves has been a hallmark of infectious disease outbreaks. Repeated surges in infections pose significant challenges to public health systems, yet the mechanisms that drive these waves remain insufficiently understood. Most prior models attribute epidemic waves to exogenous factors, such as transmission seasonality, viral mutations, or implementation of public health interventions. We show that epidemic waves can emerge autonomously from the feedback loop between infection dynamics and human behavior. Our results are based on a behavioral framework in which individuals continuously adjust their level of risk mitigation subject to their perceived risk of infection, which depends on information availability and disease severity. We show that delayed behavioral responses alone can lead to the emergence of multiple epidemic waves. The magnitude and frequency of these waves depend on the interplay between behavioral factors (delay, severity, and sensitivity of responses) and disease factors (transmission and recovery rates). Notably, if the response is either too prompt or excessively delayed, multiple waves cannot emerge. Our results further align with previous observations that adaptive human behavior can produce non-monotonic final epidemic sizes, shaped by the trade-offs between various biological and behavioral factors–namely, risk sensitivity, response stringency, and disease generation time. Interestingly, we found that the minimal final epidemic size occurs on regimes that exhibit a few damped oscillations. Altogether, our results emphasize the importance of integrating social and operational factors into infectious disease models, in order to capture the joint evolution of adaptive behavioral responses and epidemic dynamics.

**Significance statement:** We develop a behavioral-epidemiological framework in which individuals adjust their level of risk mitigation (e.g., social distancing, mask-wearing) based on both the available information and their perceived risk of infection. We show that the feedback loop between infectious disease dynamics and human behavior can autonomously produce multiple epidemic waves. The disease dynamics are strongly influenced by the interplay between the timing, severity and sensitivity of behavioral responses, as well as transmission and recovery rates. Moreover, our results confirm that adaptive human behavior can produce non-monotonic final epidemic sizes, which we show is due to oscillatory epidemic dynamics. Interestingly, we found that in the absence of interventions, the minimal final epidemic size occurs on regimes exhibiting a few damped oscillations.

## Introduction

The recurrence of epidemic waves has been a defining characteristic of various infectious disease outbreaks throughout history. Notable examples of epidemics exhibiting multi-wave dynamics include the 1918 H1N1 “Spanish Flu” pandemic, influenza pandemics and more recent occurrences such as the 2009 H1N1 pandemic and the COVID-19 pandemic [1–3]. Repeated surges in new infections pose significant challenges to public health systems, calling for a deeper understanding of the underlying mechanisms driving such contagion waves. A key question persists: what causes the emergence of multiple waves during epidemics, and how can these waves be predicted and mitigated?

Compartmental models have been foundational in the study of infectious disease dynamics [4, 5], and numerous modifications have been introduced in an effort to understand and predict multi-wave dynamics. There is an extensive literature on epidemiological models that exhibit oscillatory dynamics. Some models emphasize the impact of biological factors, such as seasonal transmissibility, human immune response heterogeneity, spatial scale, population mobility, and viral mutation, in driving multi-wave epidemics [6–12]. Nonetheless, recent pandemics highlighted the shortcomings of these models, demonstrating that transmission dynamics both drive and are driven by individuals’ behavioral responses. Behaviors, including social distancing, mask-wearing, and changes in mobility, dynamically evolve in response to perceived infection risk, media coverage, and public health policies [12–17].

Recent studies using disease-behavior interaction models have shown that social dynamics can also induce oscillatory and even chaotic epidemic dynamics. Examples of social factors driving such dynamics include the ‘stickiness effect’ (resistance to behavioral changes) in compliance with nonpharmaceutical interventions (NPIs), early relaxation of control measures, and pandemic fatigue [18–27]. Numerous studies incorporate human behavior driven by awareness, economic incentives, and risk factors, which act at both the individual and population level [28–31]. Game theoretic approaches are also commonly used to incorporate individual behavioral choices [32–37]. These modeling approaches aim to capture the coevolution of the epidemic process and behavioral adaptations, usually assuming availability of complete, accurate and immediate information. For instance, depending on the severity of an infectious disease outbreak, people make behavioral decisions about how strictly they adhere to mask-wearing guidelines, mobility or meeting restrictions. These choices influence the spread of infection, affecting the success of interventions and even altering the trajectory of epidemics [38–41].

Despite improvement on understanding the complex dynamics between human behavioral responses during epidemics, less attention has been given to modeling the impact of information delays on the decision-making process. Some models assume that disease awareness simultaneously spreads over the population as a dual social contagion, which implicitly leads to heterogeneous behavioral responses [19, 42, 43]. However, the accuracy and availability of information depend on the identification of transmission through epidemiological monitoring systems, which may face operational constraints, such as limited resources [44, 45]. Delayed behavioral responses are well-documented in epidemiological studies. The delays may fluctuate due to a number of social and operational factors. For instance, individuals often take time to perceive the severity of an outbreak and adjust their behaviors accordingly [46–48]. On the other side, limited surveillance systems or misinformation may jeopardize individuals’ behavioral choices [45, 49].

Together, the intertwined dynamics between information, behavioral changes and disease transmission, create a series of feedback loops that shape infectious disease dynamics. Consequently, understanding the joint dynamics of behavioral adaptations and disease transmission requires to unveil the role of information availability, as behavioral-driven waves can emerge from endogenous incentives without the need for exogenous shocks. In this study, we use a behavioral-epidemiological model to examine the trade-off between disease progression, information delays, and the stringency of behavioral responses in generating oscillatory dynamics. Our modeling approach builds on the classical awareness-based models by incorporating a lagged response of the population to the infection prevalence [50–52]. We model behavioral changes as adjustments in social interactions, which ultimately affect the population’s likelihood of infection. That is, individuals choose their level of daily social interactions, given their understanding of infection risks driven by information availability. In this way, behavioral responses depend on the current or the recent state of the epidemic.

Our findings show that behavioral responses driven by immediate information reduce the peak size relative to the standard, “behavior-free” model, and avoid oscillatory dynamics. On the other hand, we show that delayed information can produce multi-wave dynamics, where the number and intensity of the waves are modulated by the trade-off between the behavioral response stringency, the information delay, and the disease generation time. Moreover, delayed behavioral responses can produce non-monotonic final epidemic sizes. The minimal final epidemic size occurs during information-behavioral regimes that produce a few damped epidemic waves. In other words, our results suggest that neither single peak scenarios nor sustained multi-wave dynamics minimize the final epidemic size.

## Methods

In this section, we present how a standard SIR-type infectious disease model can be adapted to account for average, population-wide behavioral adaptations that depend on the disease prevalence. The model is designed to capture the dynamic interplay between infection spread and collective behavior, highlighting the potential for such reactions to influence the progression and recurrence of waves during an epidemic.

In this study, we envision contact reduction as the set of behavioral changes aimed at lowering the effective transmission of the disease. Individuals’ behavioral responses represent actions to minimize their exposure risk. For instance for respiratory infectious diseases, these include practicing social distancing, wearing masks, and increasing hygiene practices. Unlike standard SIR models where the contact rate remains fixed, our model accounts for dynamic adjustments based on perceived infection risk. This adaptive behavior directly modifies the transmission rate based on the available–possibly delayed– information about the prevalence of infection. This nuanced representation of behavioral changes allows us to simulate real-world scenarios where the timing and intensity of behavioral responses play a critical role in determining the trajectory and potential waves of epidemics.

### Model description

The standard SIR model divides the population into three key compartments: susceptible (*S*), infected (*I*), and recovered (*R*). The progression of the epidemic is modeled using differential equations that describe the processes of infection and recovery. To capture the influence of behavioral responses during an outbreak, we extend this model by incorporating a population-wide contact reduction factor, driven by perceived risk. Let *r*(*I/N*) ∈ [0, 1] represent the average population-wide behavioral response. While individual risk perceptions vary, it is reasonable to assume that, at the population level, risk mitigation increases as the number of infected individuals rises, i.e., *dr/dI >* 0. This leads to the following system of (delay) differential equations, which forms the basis of our analysis:

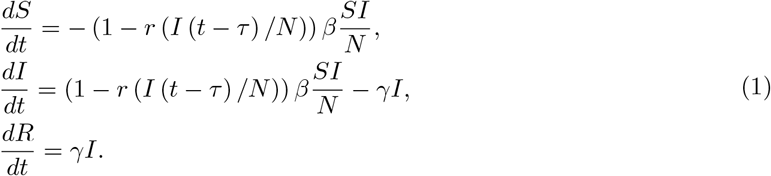

Here, *β* is the transmission rate, *γ* is the recovery rate, and *N* (= *S* + *I* + *R*) represents the total population. Note that, by design, the model dynamics do not depend on the choice of N. The contact reduction factor, *r*(*I*(*t* − *τ*)*/N*), evolves over time in response to the prevalence level (i.e., the number of active cases), with *τ* capturing the delay in data reporting and the population’s decision-making process. Since the specific form of the response function is unknown and varies based on a pathogen’s perceived risk, we explore two functional forms, each defined by two key parameters:

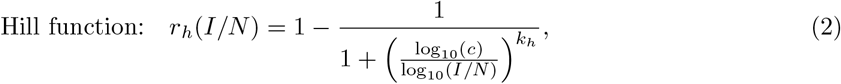

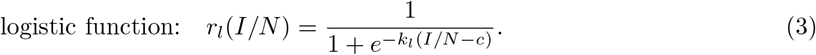

In these equations, *c* represents the prevalence threshold at which contacts are reduced by exactly 50%, and we refer to this parameter as the “behavioral response midpoint”. The parameters *k*_*h*_, *k*_*l*_ govern the sensitivity of the behavioral response, modulating how quickly the adaptation occurs as the number of cases increases. Figure 1A illustrates examples of both Hill and logistic functions. When the prevalence of the disease is low, individuals remain unaware of the outbreak, and contact reduction is minimal (i.e., *r*(*I*) ≈ 0 for small *I*). As the prevalence increases, contact reduction eventually approaches 100%, akin to a complete lockdown. Due to the logarithmic scaling in the Hill function, contact reduction increases more gradually at higher case numbers compared to the logistic function. As shown in Figure 1A, when the behavioral response midpoint is set to *c* = 2%, a Hill function with *k*_*h*_ = 16 closely matches a logistic function with *k*_*l*_ = 250 at prevalence levels around 1.5-2%, while a Hill function with *k*_*h*_ = 24 better matches the same logistic function at higher prevalence levels. This is due to the different scaling in the Hill (log-scaled) and logistic (linear-scaled) function.

**Figure 1.**
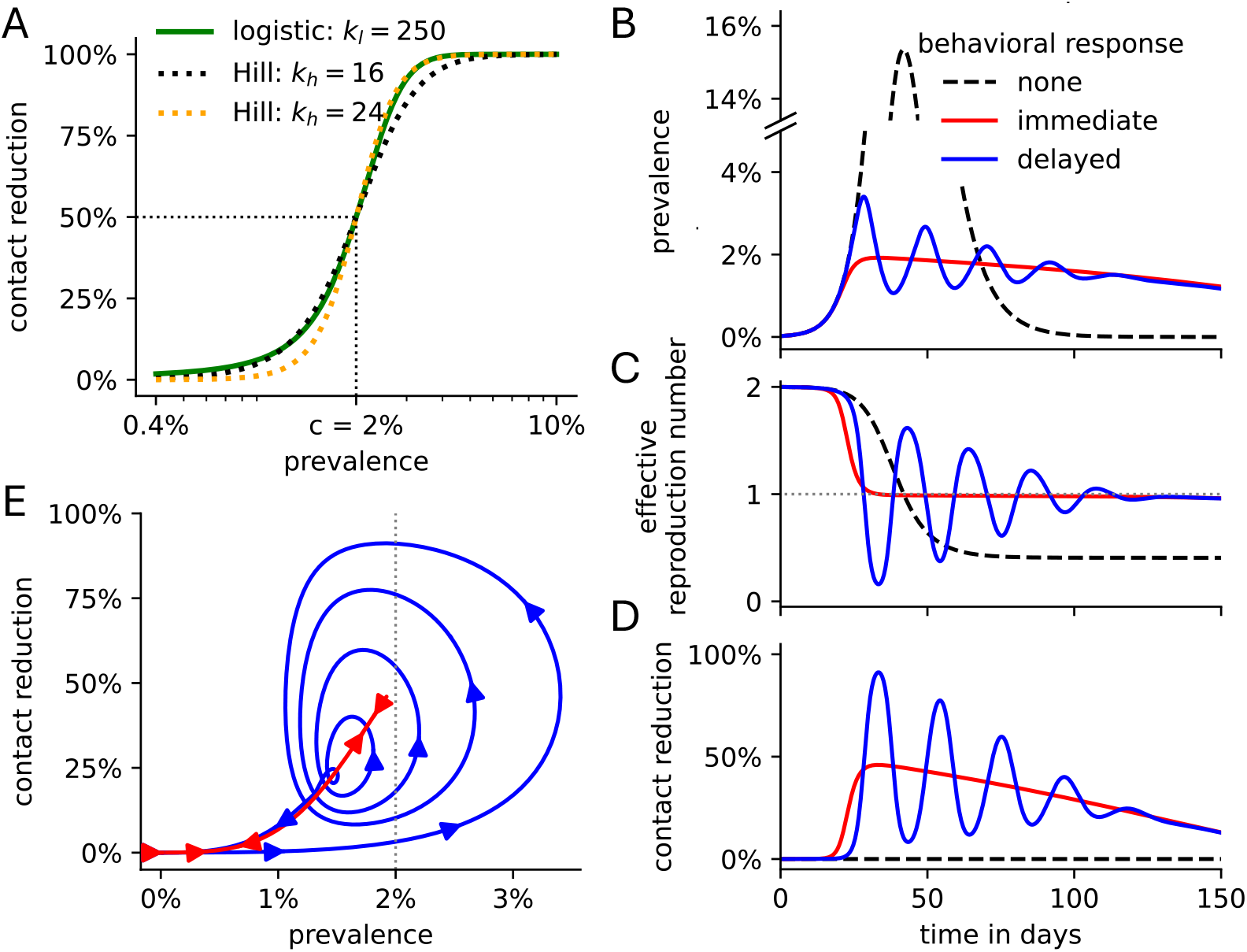
Delayed behavioral responses can induce epidemic waves. (A) Linear-scaled logistic functions (solid green line) and log-scaled Hill functions (dotted lines) can both describe the disease prevalence-dependent behavioral response. The behavioral response midpoint for all exemplary functions is fixed at *c* = 2%, while the sensitivity parameter (*k*_*h*_, *k*_*l*_, respectively) varies. (B) Disease prevalence, (C) effective reproduction number, and (D) contact reduction over time, and for different behavioral responses: none as in the standard SIR model (dashed black lines), immediate (*τ* = 0; solid red lines), delayed (*τ* = 5; solid blue lines). (E) Trajectory of the prevalence and contact reduction under an immediate (red) and delayed (blue) behavioral response. The arrows indicate the direction of the change over time. (B-E) All non-specified parameters are at their default values listed in Table 1. Specifically, *c* = 2% and *k*_*h*_ = 16.

### Effective reproduction number

The effective reproduction number, *R*_eff_(*t*), quantifies the expected number of secondary infections caused by an infected individual at a specific time *t* [53, 54]. *Unlike the basic reproduction number, ℛ*_*0*_ *= β/γ*, which assumes that the entire population is susceptible (except for an arbitrarily small number of initially infected individuals), *R*_eff_(*t*) varies over the course of an outbreak. This variation occurs due to factors such as the depletion of the susceptible population or behavioral changes in response to perceived risk. In this study, *R*_eff_(*t*) plays a key role in explaining the impact of incorporating information delays, which leads to the emergence of epidemic waves driven by changes in population-wide contact levels as the disease prevalence fluctuates. The rate of change in the number of infected individuals can be expressed as

**Table 1.**
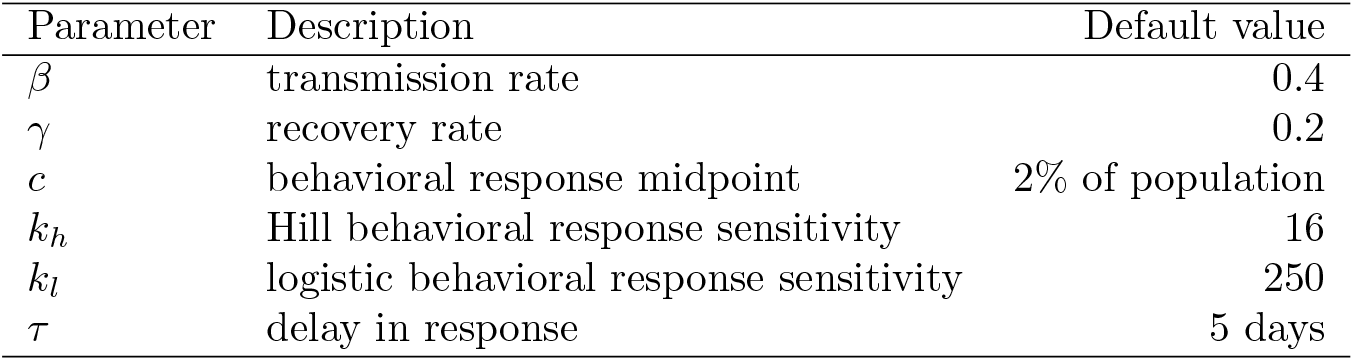
Model parameters.

| Parameter | Description | Default value |
| --- | --- | --- |
| $\beta$ | transmission rate | 0.4 |
| $\gamma$ | recovery rate | 0.2 |
| $c$ | behavioral response midpoint | 2% of population |
| $k_h$ | Hill behavioral response sensitivity | 16 |
| $k_l$ | logistic behavioral response sensitivity | 250 |
| $\tau$ | delay in response | 5 days |

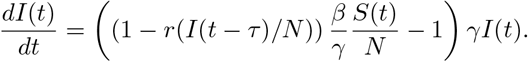

The effective reproduction number with information delay *τ* is then given by

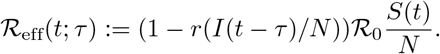

Note that the disease prevalence increases (i.e., ^*dI*(*t*)^ > 0) if and only if ℛ_eff_(*t*; *τ*) > 1.

### Simulation

We employed the fourth-order Runge-Kutta method (RK4) to simulate the model dynamics with a time step of Δ*t* = 0.1 [55, 56]. The RK4 method provides a computationally efficient approach for solving ordinary differential equations (ODEs) by evaluating the derivatives at intermediate points between time steps and taking a weighted average of these derivatives. The use of the high-performance Python compiler Numba substantially improved the compute time [57]. To account for a delay of *τ* in the reporting of cases and subsequent decision-making, we track the history of the number of infected individuals *I* over time in the array *I*_history_. The values of *I*_history_ represent the number of infected individuals at previous time points, which is necessary for simulating delayed effects on the response function in the model. To ensure the simulation starts with a consistent history, the initial values of *I*_history_ are all set to the initial number of infections, *I*(0). Throughout, we used *I*(0) = 0.02%. That is,

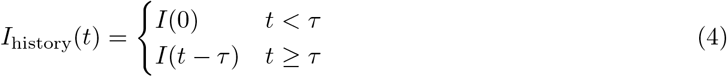

This history tracking method enables an accurate modeling of delays without introducing substantial computational cost. Table 1 describes all model parameters and their default values that are used throughout unless otherwise stated. All simulations were conducted using Python 3.11.5.

### Counting waves in disease dynamics

To quantify the number of waves in the model dynamics, we define a wave as a significant peak (i.e., local maximum) in the number of infected individuals over time. We counted peaks using the `find_peaks` algorithm from the Python library `scipy.signal`. Each peak possesses a prominence value, which quantifies the least drop in height necessary in order to get from the peak to any point with even higher value. We used a prominence threshold of 0.2% to ensure that only notable peaks in the number of infected individuals are counted as independent waves, filtering out minor fluctuations. The total number of waves is then defined as the number of peaks in the prevalence function over time. Note that the minimal number of waves is one, as long as the number of initially infected individuals is greater than the prominence threshold of 0.2%.

## Results

### Immediate behavioral adjustment in response to an infectious disease outbreak

The standard SIR model (Eq. 1 with *r* ≡ 0) possesses two parameters: the transmission rate *β* and the recovery rate *γ*. From these parameters, we can derive the basic reproduction number ℛ_0_ = *β/γ*, which describes the expected number of secondary infections caused by the first infected person when everyone else is still susceptible. Here, we assume *β* = 0.4, *γ* = 0.2 so that ℛ_0_ = 2. Since ℛ_0_ > 1, the number of infected individuals increases over time until the number of susceptibles has been depleted by 1*/*ℛ_0_, corresponding to ℛ_eff_(*t*) = 1 (Fig. 1B,C). Beyond this peak, the disease prevalence decreases. While ℛ_0_ =ℛ_eff_(0) is constant, the effective reproduction number ℛ_eff_(*t*) decreases over time as the number of remaining susceptibles declines (Fig. 1C). This yields a single, prominent epidemic peak.

In reality, individuals decrease their effective contacts (through social distancing, mask wearing, etc.) in response to a severe infectious disease outbreak, as exemplified by the recent COVID-19 pandemic [58]. Aggregated individual-level behavior gives rise to a population-wide effective contact reduction, which depends on the current or recent level of disease prevalence and can be qualitatively captured by both Hill functions (Eq. 2) and logistic functions (Eq. 3; Fig. 1A). Prior to awareness and media attention, a population does not engage in outbreak-related risk mitigation measures (i.e., *r*(*I/N* = 0) = 0). As the prevalence of an infectious disease rises, an increasing number of individuals fear getting infected, and more risk mitigation policies are put in place (i.e., *dr/dI >* 0), both at the individual and the societal level. We hypothesized, in the absence of data, that the population-wide reduction in effective contacts likely follows a logarithmic scale, which means that a change in prevalence from, e.g., 1% to 2% would result in the same change in behavioral response as a change from 2% to 4%. Accordingly, we used a log-scaled Hill function to model this response in our main results. For comparison, results based on a linear-scaled logistic function, which yielded qualitatively similar outcomes (Fig. S1 and Fig. S2), are presented in the supplement.

The Hill functional response is characterized by two parameters: the behavioral response midpoint *c*, at which contacts are reduced by exactly 50%, and the parameter *k*_*h*_, which describes the sensitivity of the behavioral response to changes in disease prevalence. In the absence of data, we fixed *c* = 2% and *k*_*h*_ = 16 and varied these parameters in later sensitivity analyses. In reality, these parameters will depend on the severity of the disease. For example, people will engage in higher levels of risk mitigation (i.e., *c* is lower) during an Ebola outbreak (characterized by high hospitalization and mortality rates) versus a seasonal flu outbreak. In the scenario where contact levels depend on current disease prevalence (i.e., no delay (*τ* = 0) in case-reporting and decision-making), disease dynamics differ substantially from standard SIR dynamics: the effective reproduction number decreases to 1 much faster – before 1*/ℛ* _0_ of individuals have become infected (Fig. 1B,C). This is due to the prevalence-dependent reduction in effective contacts, driven by the immediate and sustained transmission-reducing behavioral adaptation (Fig. 1D). The effective reproduction number then stabilizes for an extended period of time at values just below 1. During this period, the overall activity level of the population gradually increases, while the disease prevalence and the number of susceptible individuals both steadily but slowly decline. Eventually, ℛ_eff_ drops markedly below 1, quickly leading to an end of the outbreak. While the shape of the epidemic curve is very different, an immediate contact reduction (i.e., *τ* = 0) only yields one, albeit prolonged epidemic wave.

### Delayed behavioral adjustment in response to an infectious disease outbreak

We next investigated the effect of delay in behavior adjustment on the shape of epidemic curves. In reality, the delay is always positive because information on new infections first requires diagnosis and then reporting. The detrimental impact of delays in diagnosis on individual disease progression, disease spread, and economic outcomes has been extensively studied for many infectious diseases, e.g., COVID-19 [46], African viral hemorrhagic fever [47] and foot-and-mouth disease [59]. Here, we explore the effect of delays on inducing epidemic waves. Assuming a constant delay of *τ* = 5 days, the initial outbreak size increases quickly due to the unawareness of the population. Once contacts are reduced in response to the large outbreak, the effective reproduction number drops quickly below 1 giving rise to a first peak in disease prevalence (Fig. 1B-D). Following the drop in prevalence, the population-wide activity level increases again after a delay of *τ* = 5 days. This rise leads to ℛ_eff_ > 1 and the emergence of a second epidemic peak, which is less prominent than the first due to the reduced number of remaining susceptible individuals. This pattern repeats a few more times, with each subsequent peak exhibiting a smaller amplitude in prevalence (Fig. 1E). Eventually, the effective reproduction number stabilizes just below 1. From this point forward, disease prevalence gradually declines, resembling the trend observed in the absence of a delay. The shape of the epidemic curve depends strongly on the delay parameter. When the delay is short (e.g., *τ* = 2 days), the disease dynamics resembles the case of no delay, characterized by a single, prolonged low-prevalence epidemic (Fig. 2A). After population-wide effective contacts are reduced by about 1*/ℛ*_0_, activity levels begin to slowly increase as the prevalence level decreases (Fig. 2B). On the other hand, when the delay is very long (e.g., *τ* = 18 days), the disease dynamics resembles the standard SIR model, characterized by one high prevalence peak. With long delays, the population-wide behavior adjustment starts too late during the outbreak and can only slightly lower peak prevalence levels.

**Figure 2.**
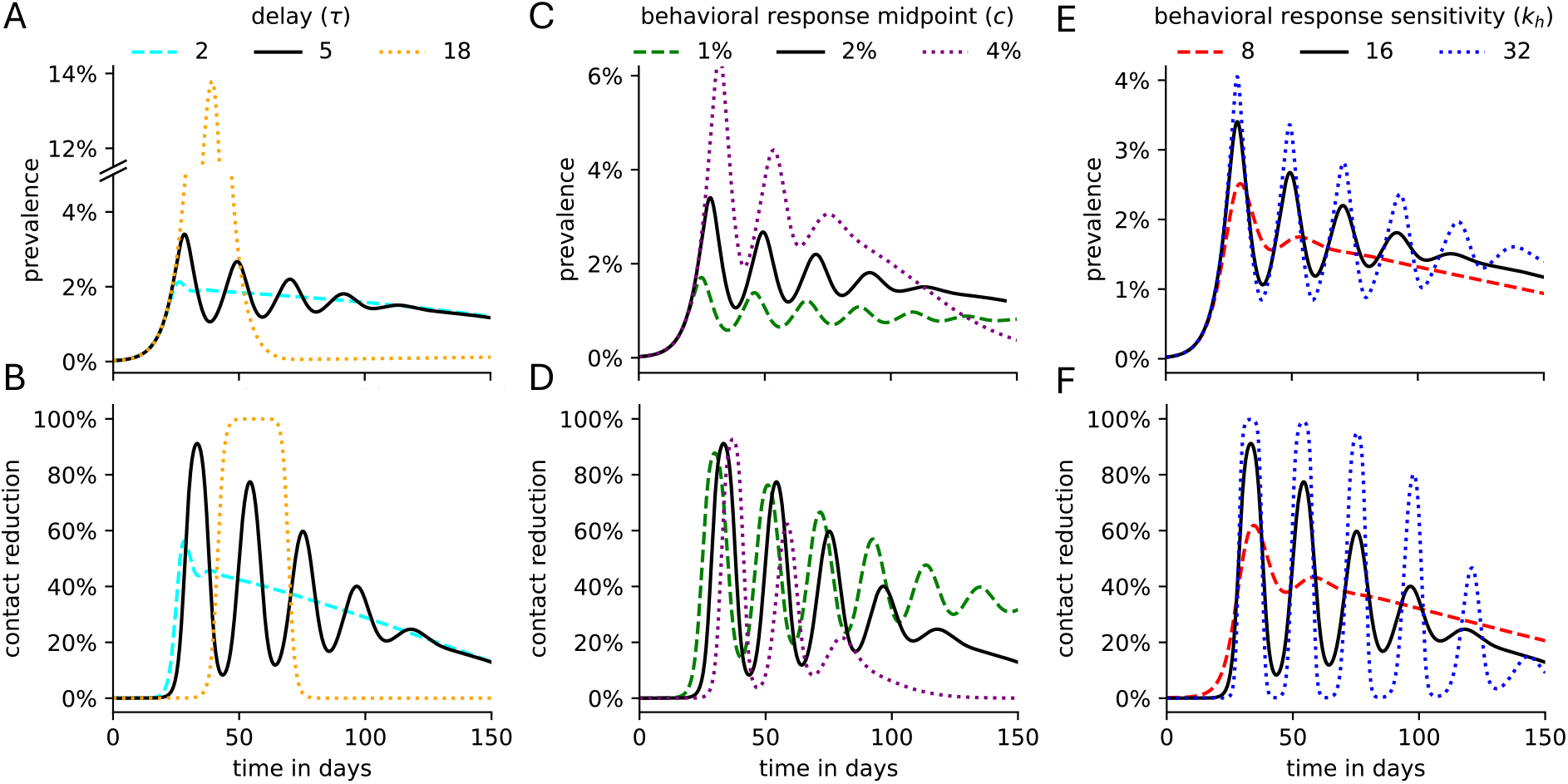
Disease dynamics and population-wide contact reduction for a variety of delays and Hill response functions. Given a delay of *τ* days and a population-wide contact reduction function, parametrized by the behavioral response midpoint *c* and the sensitivity *k*_*h*_, the (A,C,E) disease prevalence and (B,D,F) population-wide contact reduction is plotted over time for several (A,B) *τ* -values, (C,D) *c*-values and (E,F) *k*_*h*_-values. All non-specified parameters are at their default values listed in Table 1. In all sub panels, the solid black line depicts the dynamics for *τ* = 5, *c* = 2%, *k*_*h*_ = 16.

We explore the impact of varying the response function shape (parametrized by the behavioral response midpoint *c* and the sensitivity parameter *k*_*h*_), to represent diverse expected population-wide behavioral response. Higher *c*-values imply that contacts are reduced less strongly, leading to a larger first epidemic peak (Fig. 2C,D). This increased outbreak causes a larger reduction in the number of susceptibles, which explains why higher *c*-values are associated with fewer epidemic peaks and disease prevalence that begins more rapidly to drop steadily towards zero, as in the case of no delay (Fig. 1B-E). If the contact reduction is less sensitive to the prevalence level (i.e., low *k*_*h*_-values), the contact reduction begins at lower prevalence levels (see Fig. 1A), leading to a lower first epidemic peak (Fig. 2E,F). The lower sensitivity also implies that the level of contact reduction does not change dramatically as the first wave of infections declines, yielding just one more faint peak in prevalence numbers. On the contrary, high *k*_*h*_-values (e.g., *k*_*h*_ = 32) imply nearly complete lockdowns and relaxations between each epidemic wave, characterized by close to 100% and 0% population-wide effective contact reduction, respectively. A more sensitive behavioral response function (i.e., high *k*_*h*_-values) induces more epidemic waves. This cannot be explained by variation in the number of susceptibles, which declines basically at the same speed for all *k*_*h*_-values (indicated by the comparable area under the prevalence curves in Fig. 2E). Interestingly, the periodicity of the epidemic waves appears to solely depend on the delay parameter *τ* but not on the midpoint or the sensitivity of the behavioral response function.

To further explore the connection between the number of epidemic waves and parameter choices, we varied the delay *τ* between 0 and 20 days in addition to one of the model parameters: behavioral response midpoint *c*, behavioral response sensitivity *k*_*h*_, transmission rate *β*, and recovery rate *γ* (Fig. 3). These two-dimensional sensitivity analyses expand the previous findings. Whenever the delay is very small, there exists only one wave, as seen in Fig. 1B-E for the boundary case of *τ* = 0. Irrespective of the specific delay, lower *c*-values generally induce more waves, which can be explained by the earlier onset of behavioral response and subsequent smaller reduction in susceptibles per wave (Fig. 3A). For a fixed behavioral response function (i.e., fixed *c*), most waves occur at a delay of 5-7 days, with the wave-maximizing delay decreasing slowly as *c* increases. A more sensitive behavioral response function generally yields more waves (Fig. 3B). The wave-maximizing delay depends strongly on the behavioral response sensitivity. At high *k*_*h*_-values (e.g., *k*_*h*_ = 36), a delay of 2.25 days suffices to induce nine waves, while this delay causes only a single wave when *k*_*h*_ *<* 14.3. At lower *k*_*h*_-values, a longer delay is required for multiple waves to emerge.

**Figure 3.**
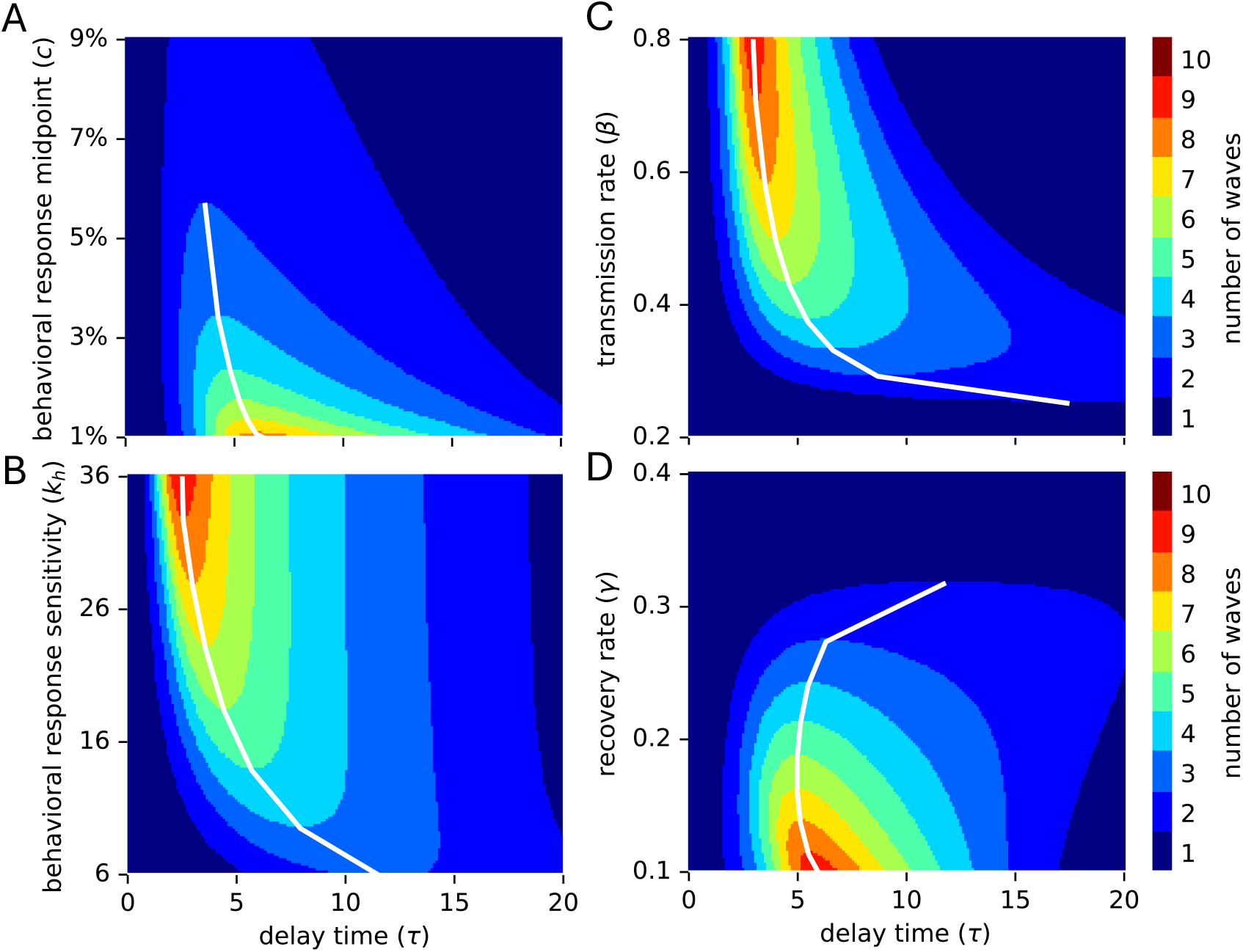
Two-dimensional sensitivity analysis. The number of epidemic waves is shown for a range of values for the delay parameter (*τ*, x-axis) and another model parameter (y-axis): (A) behavioral response midpoint *c*, (B) behavioral response sensitivity *k*_*h*_, (C) transmission rate *β*, (D) recovery rate *γ*. White lines connect the highest (in A,D) or lowest (in B,C) model parameter value and associated delay value that yields a specific number of multiple waves. All non-specified parameters are at their default values listed in Table 1.

### Occurrence of epidemic waves depends on the interplay between delay in behavior adjustment and disease generation time

In all results thus far, the basic reproduction number ℛ_0_ = *β/γ* was 2, assuming a transmission rate of *β* = 0.4 and recovery rate *γ* = 0.2. At higher transmission rates (and thus higher reproduction numbers), the number of waves increases and the wave-maximizing delay decreases (Fig. 3C). Similarly, when assuming slower recovery rates (and thus higher reproduction numbers), the number of epidemic waves increases as well (Fig. 3D). While increasing transmission rates and decreasing recovery rates both modulate the basic reproduction number in the same way, there exists a major difference between the two approaches, which is captured by the disease generation time–the average time between the infection of a person and the onward transmission by this person [60]. This key epidemiological metric (which is often approximated by the serial interval) is crucial for understanding how quickly a disease can spread within a population. Fast-spreading diseases such as COVID-19 have a short disease generation time and are characterized by comparably high transmission and recovery rates, while slow-spreading pathogens such as HIV-1 possess the opposite: long disease generation times and comparably low transmission and recovery rates (an infected person may even never naturally recover from some slow-spreading diseases). For instance, setting *β* = 0.8, *γ* = 0.2 or *β* = 0.4, *γ* = 0.1 both yields ℛ_0_ = 4. The disease generation time in the latter case is, however, twice as long. For a fixed behavioral response function, both parameter choices can give rise to a maximum of nine waves (Fig. 3C,D). If *β* = 0.8, *γ* = 0.2, this maximal number of waves occurs at a delay *τ* = 3. On the other hand, if *β* = 0.4, *γ* = 0.1, the wave-maximizing delay is exactly twice as high with *τ* = 6.

To further investigate the relationship between the disease generation time and the wave-maximizing delay, we performed a four-dimensional sensitivity analysis. We modulated a given ℛ_0_-value by a combination of transmission and recovery rates and counted the maximal number of waves and the wave-maximizing delay for four different behavioral response functions, characterized by two values for the midpoint *c* and two values for the sensitivity parameter *k*_*h*_ (Fig. 4A). Higher ℛ_0_-values generally caused more waves, which can likely be explained by the stronger initial outbreak and a subsequent stronger behavioral response, followed by waves of restriction and relaxation that decrease in amplitude. For any ℛ_0_, the maximal number of waves did not differ much when varying the disease generation time by an order of magnitude. The two parameters governing the shape of the behavioral response function exhibited the trends already observed in Fig. 2 and Fig. 3A,B: a highly sensitive behavioral response function that initiates behavior modification at low prevalence levels generally yields more waves. Irrespective of the shape of the behavioral response function, slower-spreading diseases exhibited the maximal number of waves at longer delays, providing further evidence for a strong association between the disease generation time and the wave-maximizing delay. For a fixed ℛ_0_, the wave-maximizing delay proved inversely proportional to the transmission rate (Fig. 4B) and thus also to the recovery rate. Since the disease generation time is the reciprocal of the recovery rate, the wave-maximizing delay is directly proportional to the disease generation time (Fig. 4C).

**Figure 4.**
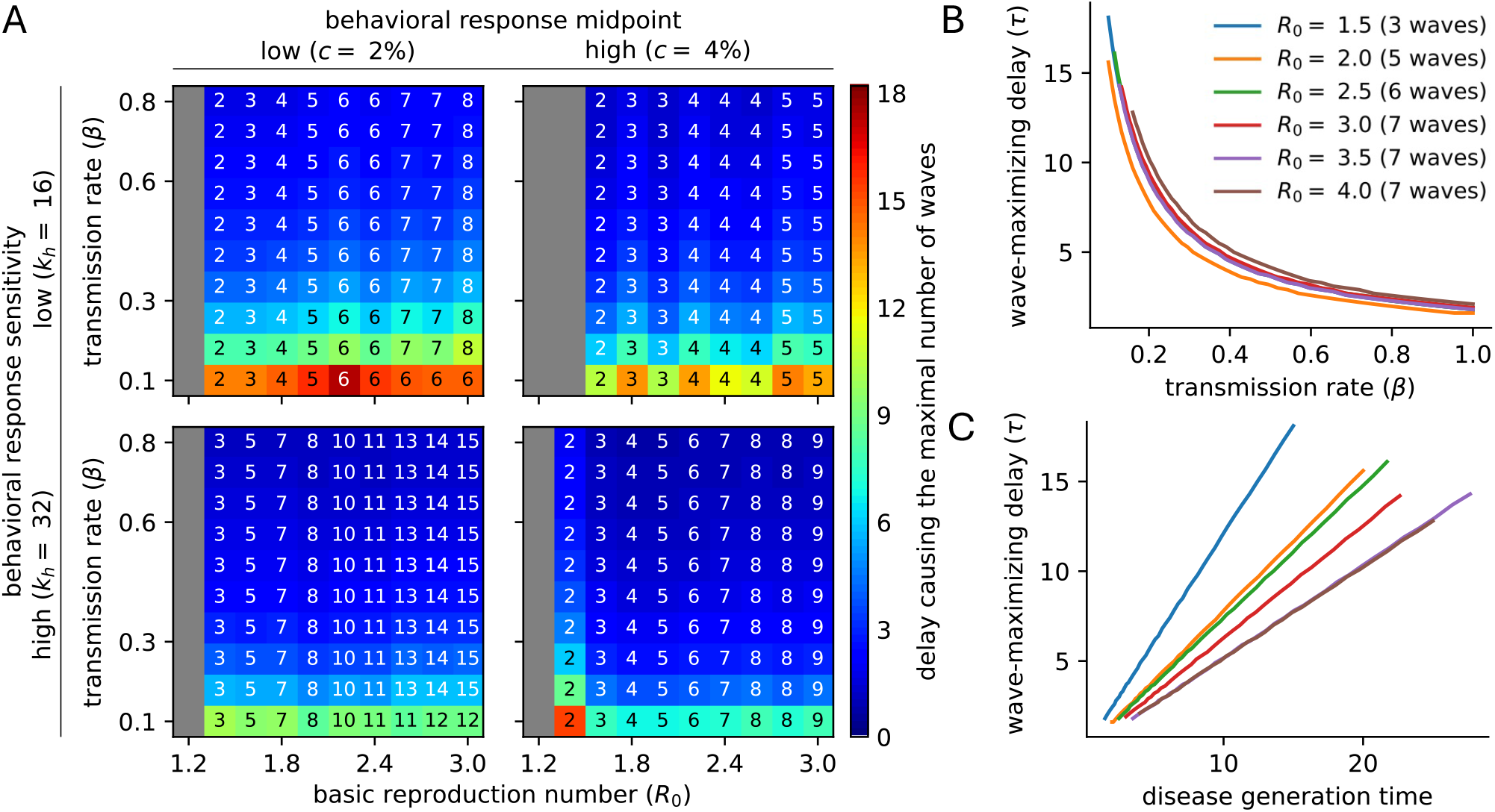
Four-dimensional sensitivity analysis. (A) The delay causing the maximal number of waves (color) and the corresponding number of waves (numbers in each cell) are shown for different basic reproduction numbers ℛ_0_ (x-axis), different disease generation times modulated by the transmission rate *β* (y-axis), as well as for four different shapes of the population-wide behavioral response function, parametrized by the behavioral response midpoint *c* and the sensitivity *k*_*h*_. Gray cells indicate that the model behaves as the standard SIR model and exhibits only a single epidemic wave, irrespective of the delay parameter. (B,C) For a fixed ℛ_0_ value and fixed behavioral response function (*c* = 2%, *k*_*h*_ = 16), the wave-maximizing delay (y-axis) is inversely proportional to (B) the transmission rate *β* and thus directly proportional to (C) the disease generation time. For each ℛ_0_ value, the line only extends across those x-values that yield the respective maximal number of waves, which is indicated in the legend in (B).

### Population-wide behavioral adjustments non-trivially affect the final epidemic size

In a standard SIR model (e.g., without reinfection and demographics), the final epidemic size describes the proportion of the total population that has been infected by the time the epidemic ends. For the standard SIR model (Eq. 1 with *r* ≡ 0), there exists a one-to-one correspondence between the final epidemic size *R*_*∞*_ and the basic reproduction number ℛ_0_, implicitly described by

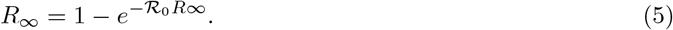

If *β* = 0.4, *γ* = 0.2, as assumed by default here, ℛ_0_ = 2 yielding *R*_*∞*_ = 79.7%. Across a wide range of delay parameters and shapes of the behavioral response function (parametrized by *c* and *k*_*h*_), the final epidemic size varied between 52% and 71% (Fig. 5A,B). This highlights that a population-wide prevalence-dependent behavioral response generally reduces *R*_*∞*_, despite resulting in potentially multiple epidemic waves. Higher behavioral response midpoints *c* and sensitivity values *k*_*h*_ are generally associated with higher *R*_*∞*_-values. However, this trend is far from monotonic. Parameter choices close to the threshold where the number of epidemic waves changes give rise to lower final epidemic sizes. Higher *c*-values or a longer delay in population-wide behavioral response both yield an initial epidemic wave that is more severe, associated with a higher peak prevalence level and more infections during the first wave (Fig. S3). The increased depletion of the pool of susceptibles can however lead to the avoidance of a second wave (if persistently ℛ_eff_ *<* 1) and thus to a final epidemic size that is lower than in the case of two smaller waves of infections.

**Figure 5.**
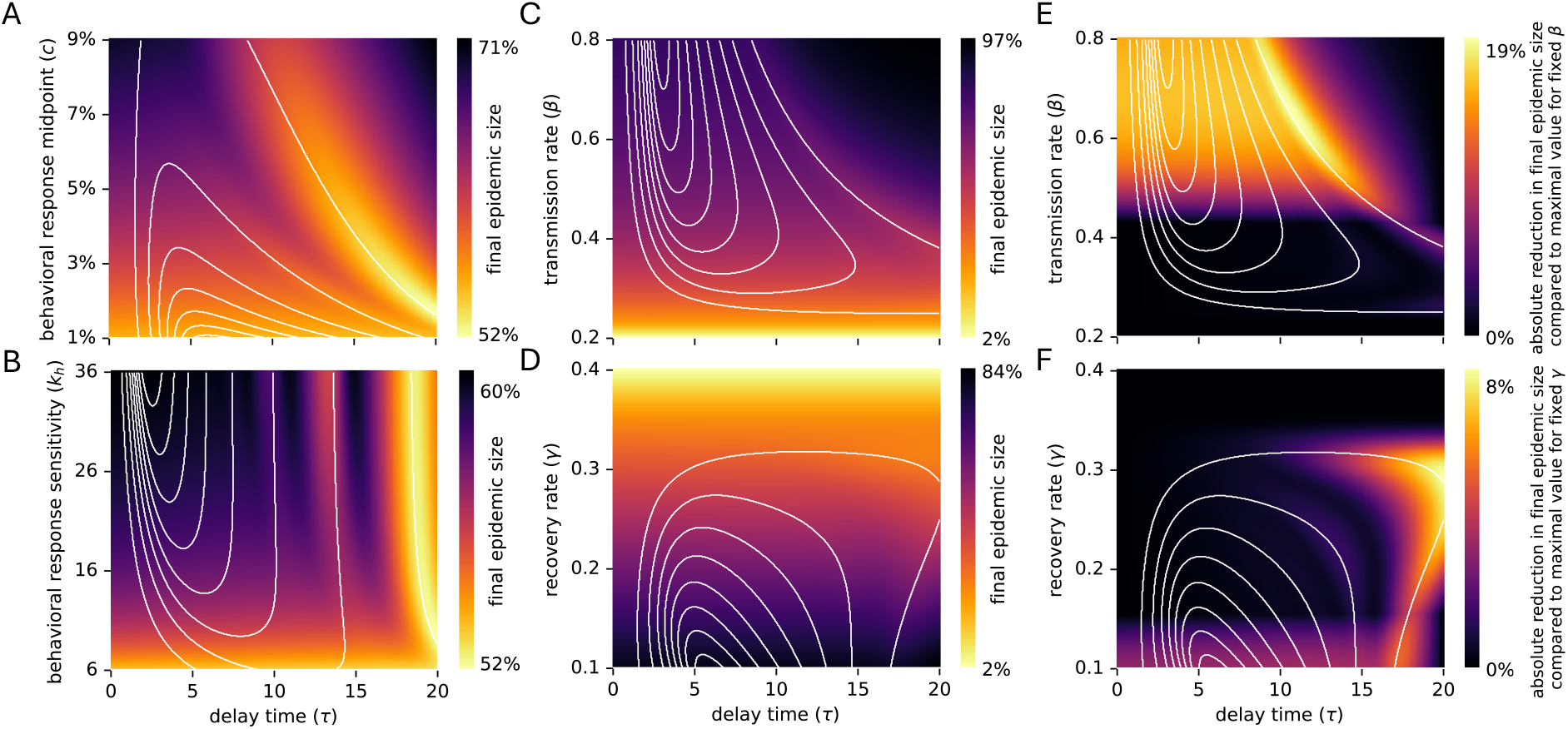
Disease and behavior-related parameters affect the final epidemic size non-monotonically. (A-D) The final epidemic size is shown for a range of values for the delay parameter (*τ*, x-axis) and another model parameter (y-axis): (A) behavioral response point *c*, (B) behavioral response sensitivity *k*_*h*_, (C) transmission rate *β*, (D) recovery rate *γ*. (E,F) The absolute reduction in final epidemic size is shown for a range of delays (x-axis) compared to the maximal value observed for a fixed (E) transmission rate or (F) recovery rate. (A-F) White lines depict the thresholds where the number of waves changes, as shown in Fig. 3. All non-specified parameters are at their default values listed in Table 1.

As expected, higher transmission rates and lower recovery rates, both associated with higher ℛ_0_-values, generally cause a larger total number of infections over the course of the epidemic (Fig. 5C,D). However, the just-described phenomenon is also evident for variation in these parameters: Specifically at the transition from one to two waves, the final epidemic size can be substantially lower (Fig. 5E,F). In other words, when accounting for delayed population-wide behavioral adjustment, a higher basic reproduction number, modulated by higher *β* or lower *γ* values, does not necessarily result in a higher total number of infections. Instead, the final epidemic size depends on the length of the delay in case-reporting and decision-making.

## Discussion

Using a simple yet insightful behavioral-epidemiological model, we examined the impact of information delay on both the generation of oscillatory epidemic dynamics and consequently on the final epidemic size. We explored the trade-offs between different stringency levels of behavioral responses, information time lags, and pathogen characteristics (specifically, the disease generation time). Our results show that immediate risk-based behavioral adaptation effectively avoids high prevalence levels by distributing infections over time. Delays in information availability and decision-making can greatly impact the shape of infectious disease dynamics. Delayed behavioral responses can induce oscillatory dynamics and produce non-monotonic final epidemic sizes. The emergence of these phenomena is modulated by the interplay between information availability, response stringency, and disease generation time. Particularly, our results show that (i) adaptive human behavior shapes the amplitude and frequency of epidemic waves; (ii) the final epidemic size exhibits non-monotonic changes as a function of several behavior or disease parameters, where the minimal final epidemic size is attained on regimes that exhibit a few damped oscillations (i.e., when the number of epidemic waves changes).

Our findings indicate that the emergence of epidemic waves is heavily influenced by the feedback between the timing, severity and sensitivity of the behavioral response, as well as transmission and recovery rates. Notably, if the response is either too prompt or excessively delayed, multiple waves do not emerge. Significantly delayed responses may come too late, missing the peak of new infections and depleting the susceptible population, resulting in fewer or no subsequent waves. Conversely, hardly delayed responses yield a prolonged, low-prevalence first wave and lower the susceptible pool before any decline in cases, preventing the formation of additional waves. Interestingly, the range of information time lags that yields multi-wave dynamics depends on the disease generation time, which proved to be directly proportional to the wave-maximizing delay.

Moreover, our results confirm previous observations by Qiu *et. al*. and Morsky *et. al*. about the non-monotonic final epidemic size [18, 19]. In contrast to these studies, we show that the incorporation of a continuous reaction space prevents discontinuities in the final epidemic size, avoiding the emergence of threshold points. It is known that the timing and intensity of behavioral responses are not uniform across populations. Variations in awareness, risk perception, age, socioeconomic status, cultural background, and adherence to protective measures contribute to a gradual and uneven shift in collective behavior [25,31,61]. Our model partly captures this variability, avoiding rigid step-wise behavioral regimes and instead allowing for smooth transitions in effective contact reduction, capturing the average population-wide behavior. It is worth to notice that our results focus on the final epidemic size in the absence of centralized interventions. Future research could consider more complex models that explore the interplay between potential centralized and decentralized interventions available to contain epidemics.

In this study, we assumed that behavioral responses are exclusively driven by the disease prevalence and do not vary due to factors such as “epidemic fatigue” or economic constraints, which would limit the frequency and action space of behavioral choices [62–65]. The recent COVID-19 pandemic highlighted that human behavior adapts over time. Epidemic fatigue was observed throughout the world, which implies that the behavioral response midpoint will likely increase over the course of an outbreak. Similarly, the delays in information availability will likely fluctuate. Delays in case-reporting will decrease as testing capacities increase. On the contrary, media coverage frequency will generally decrease, leading to potentially longer delays in risk awareness and decision-making. Further, we considered only the population-wide behavioral response, which we assumed aggregates all individual decision-making. That is, we ignored heterogeneities in compliance, risk perception, and vulnerability among different subgroups, as well as seasonality or pathogen importation/mutations [24,30,66–72]. Moreover, we assumed individuals are naive to the impact their decisions impose on others: we did not incorporate costs and benefits that behavior would have on others, missing the impact of empathy or social group affinities in structured populations.

The relative simplicity of our model enabled a comprehensive model analysis. To show that the main finding - adaptive human behavior and delays in information availability suffice for epidemic waves to emerge - is qualitatively insensitive to the specific choice of compartmental model, we analyzed the dynamics of an SEIR model, in which individuals upon infection first transition through a latency period before being infectious and counted in the disease prevalence. We found that oscillatory dynamics still emerged and the main findings were preserved (Fig. S4), although a longer latency period yielded fewer epidemic waves for a fixed delay in information availability (Fig. S5). A detailed analysis of more complicated behavioral-compartmental models constitutes an interesting avenue for future study.

The exhibited ability of epidemic waves to emerge solely due to “natural” human behavior and circumstances suggests that epidemic interventions should not only target the biological aspects of the disease but also consider the joint dynamics with the evolving behavioral responses of the population. The insights from our model could help explain recurrent patterns seen in real-world epidemics, such as early stages of the COVID-19 epidemic in the United States when behavioral responses mainly shaped transmission. Behavioral changes like social distancing and strategic contacts may independently sustain epidemic waves, highlighting the role of behavioral inertia in generating multiple peaks.

Our results demonstrate that epidemic waves can emerge autonomously from the feedback between disease dynamics and human behavior, without the need for exogenous shocks like mutations or seasonal effects. This has significant implications for public health policy and the development of integral understanding of behavioral epidemiology, as it suggests that multiple waves can occur even in the absence of any external factors. Understanding how different types of delays—whether due to social, logistical, or information factors—affect disease dynamics could refine our model and yield actionable insights for public health strategies. Our results underscore the need to integrate the interplay between behavioral and infectious disease dynamics into epidemic models, as timely and adaptive interventions could play a critical role in mitigating the impact of subsequent outbreaks. Future work to extend the developed framework would explore more complex behavioral responses, such as varying levels of compliance within subgroups of a population, or incorporating additional factors like vaccination or waning immunity. Moreover, contrasting the model to empirical epidemic data from past epidemics could help validate its predictive power and provide insights into optimizing intervention strategies to minimize the impact of future outbreaks.

In conclusion, our study fills a critical gap in the understanding of autonomous wave generation in epidemic models by linking human behavior and delays in information availability to the spread of diseases in a natural and dynamic way. By integrating behavioral responses into epidemic modeling, this work contributes to a deeper understanding of behavioral-epidemiological systems and highlights the importance of timely and sustained interventions in mitigating the effects of infectious disease outbreaks.

## Data Availability

All data produced are available online at https://github.com/ckadelka/epidemic-waves

## Acknowledgments

All code underlying this study is freely available at https://github.com/ckadelka/epidemic-waves. BE was partially supported by the NSF through DMS Award Number:2327710 and Expeditions in Computing Grant CCF-1918656. CK was partially supported by a travel grant from the Simons Foundation (grant number 712537).

## Supplementary Materials

**Figure S1.**
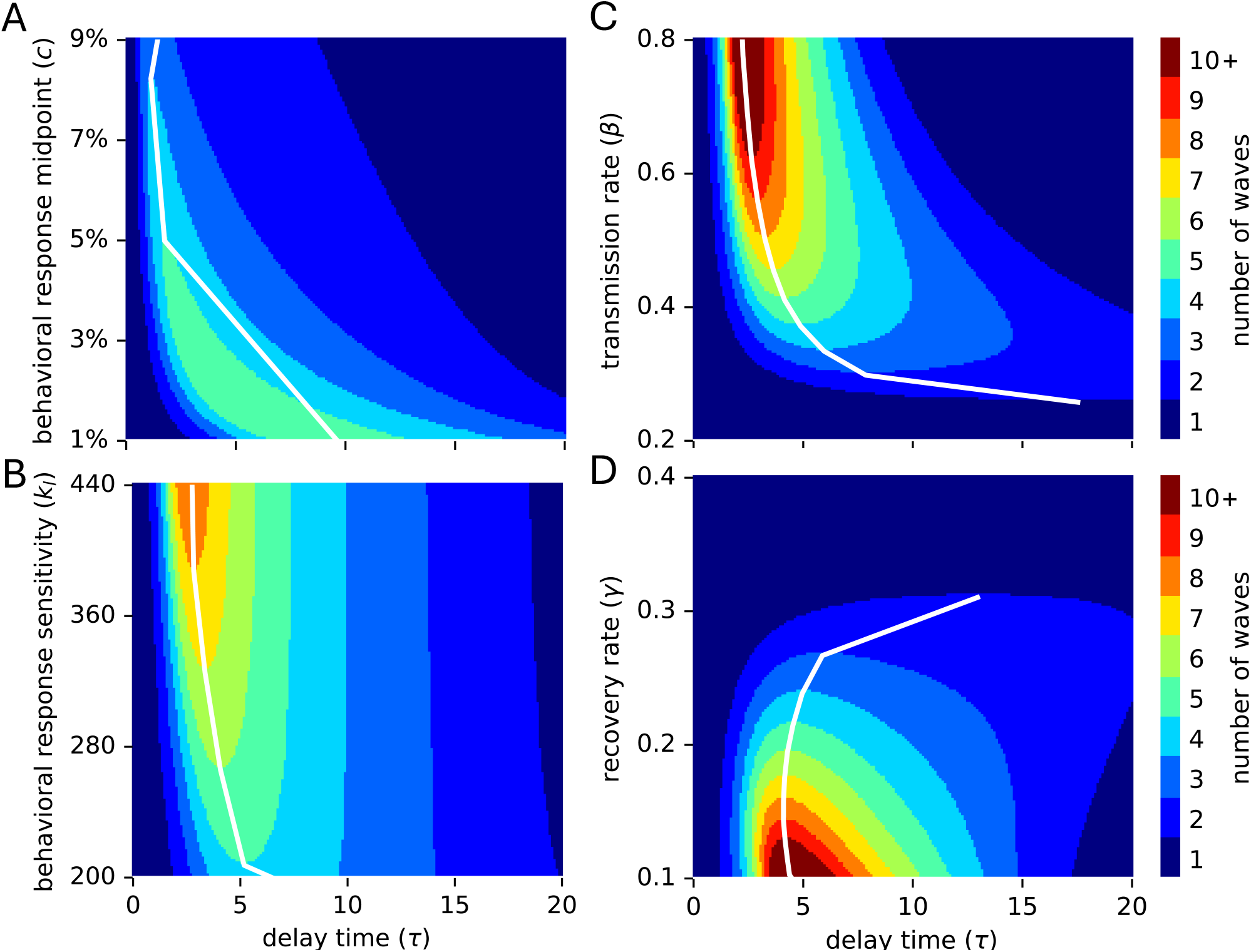
Two-dimensional sensitivity analysis, assuming a linear-scaled behavioral response function (Eq. 3). The number of epidemic waves is shown for a range of values for the delay parameter (*τ*, x-axis) and another model parameter (y-axis): (A) behavioral response midpoint *c*, (B) sensitivity of the logistic contact reduction function *k*_*l*_, (c) transmission rate *β*, (D) recovery rate *γ*. White lines connect the highest (in A,D) or lowest (in B,C) model parameter value and associated delay value that yields a specific number of multiple waves. All non-specified parameters are at their default values listed in Table 1.

**Figure S2.**
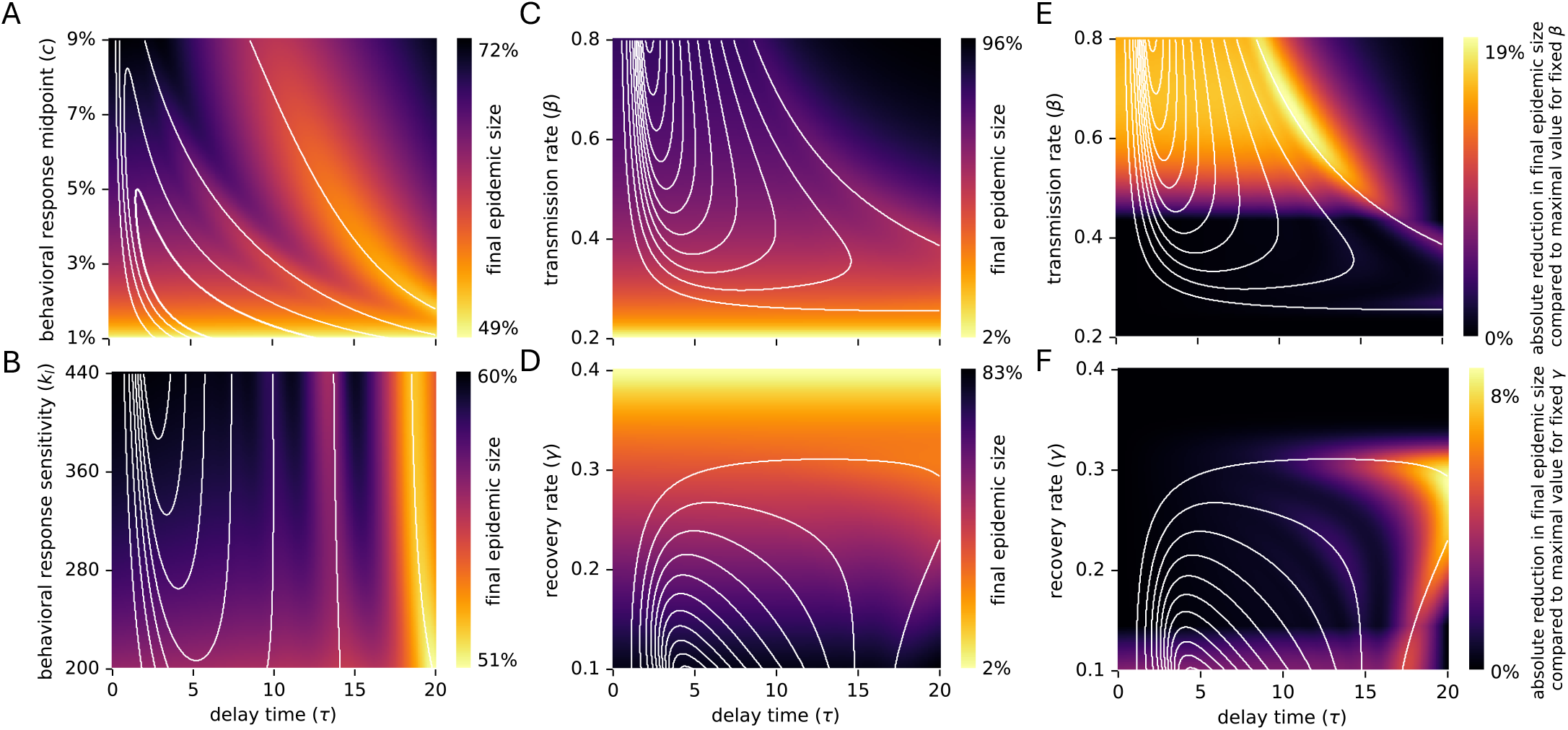
Disease and behavior-related parameters affect the final epidemic size non-monotonically, even when assuming a linear-scaled behavioral response function (Eq. 3). (A-D) The final epidemic size is shown for a range of values for the delay parameter (*τ*, x-axis) and another model parameter (y-axis): (A) behavioral response point *c*, (B) logistic behavioral response sensitivity *k*_*l*_, (C) transmission rate *β*, (D) recovery rate *γ*. (E,F) The absolute reduction in final epidemic size is shown for a range of delays (x-axis) compared to the maximal value observed for a fixed (E) transmission rate or (F) recovery rate. (A-F) White lines depict the thresholds where the number of waves changes, as shown in Fig. S1. All non-specified parameters are at their default values listed in Table 1.

**Figure S3.**
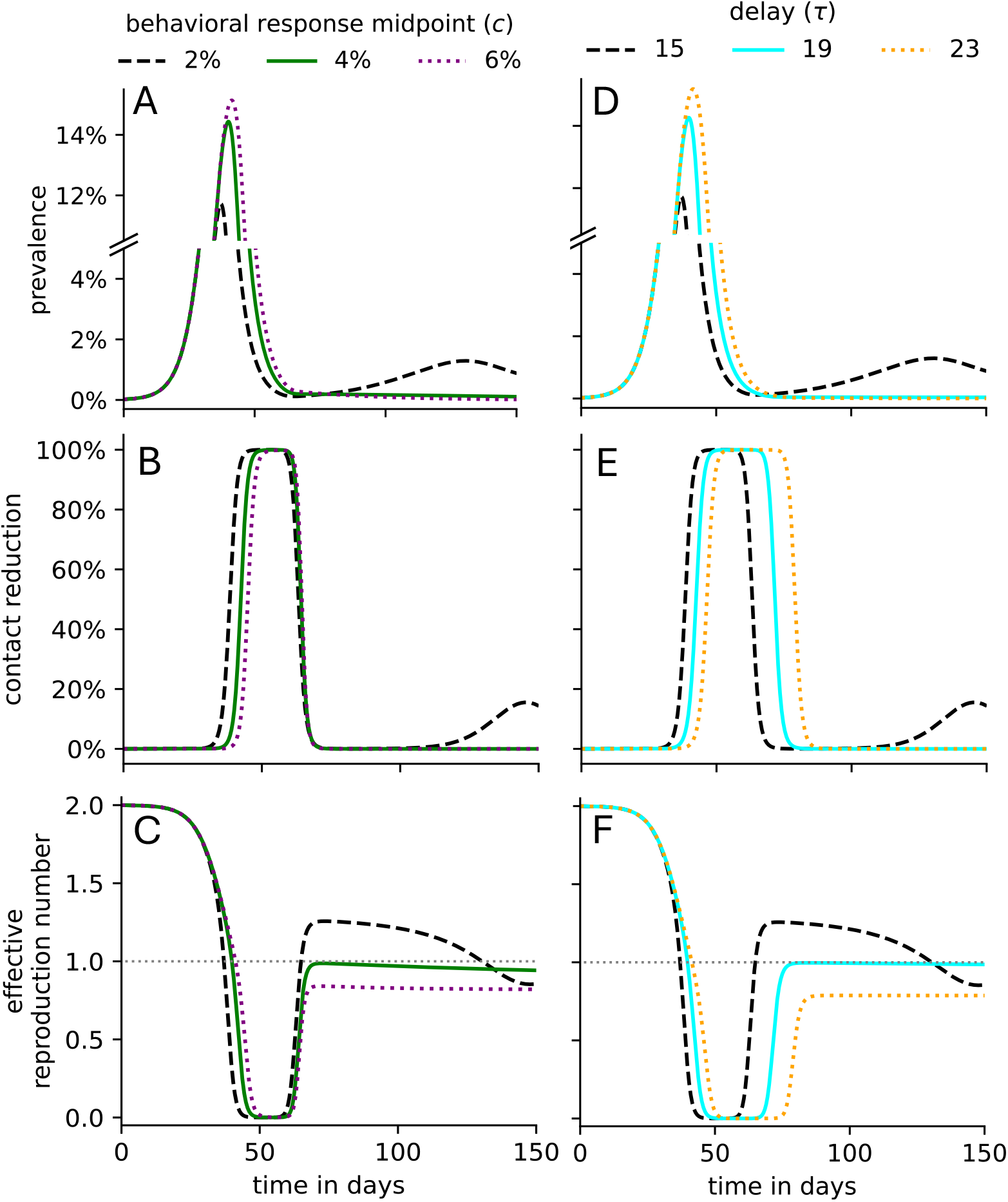
Cause of the non-monotonic final epidemic sizes. Disease dynamics and population-wide contact reduction for a variety of delays and Hill response functions. Given a delay of *τ* days and a population-wide contact reduction function, parametrized by the behavioral response midpoint *c* and the sensitivity *k*_*h*_, the (A,D) disease prevalence, (B,E) population-wide contact reduction, and (C,F) effective reproduction numbers are plotted over time for several (A-C) *c*-values (here *τ* = 15) and (D-F) *τ* -values. All non-specified parameters are at their default values listed in Table 1. In all sub panels, the dashed black line depicts the dynamics for *τ* = 15, *c* = 2%, *k*_*h*_ = 16.

**Figure S4.**
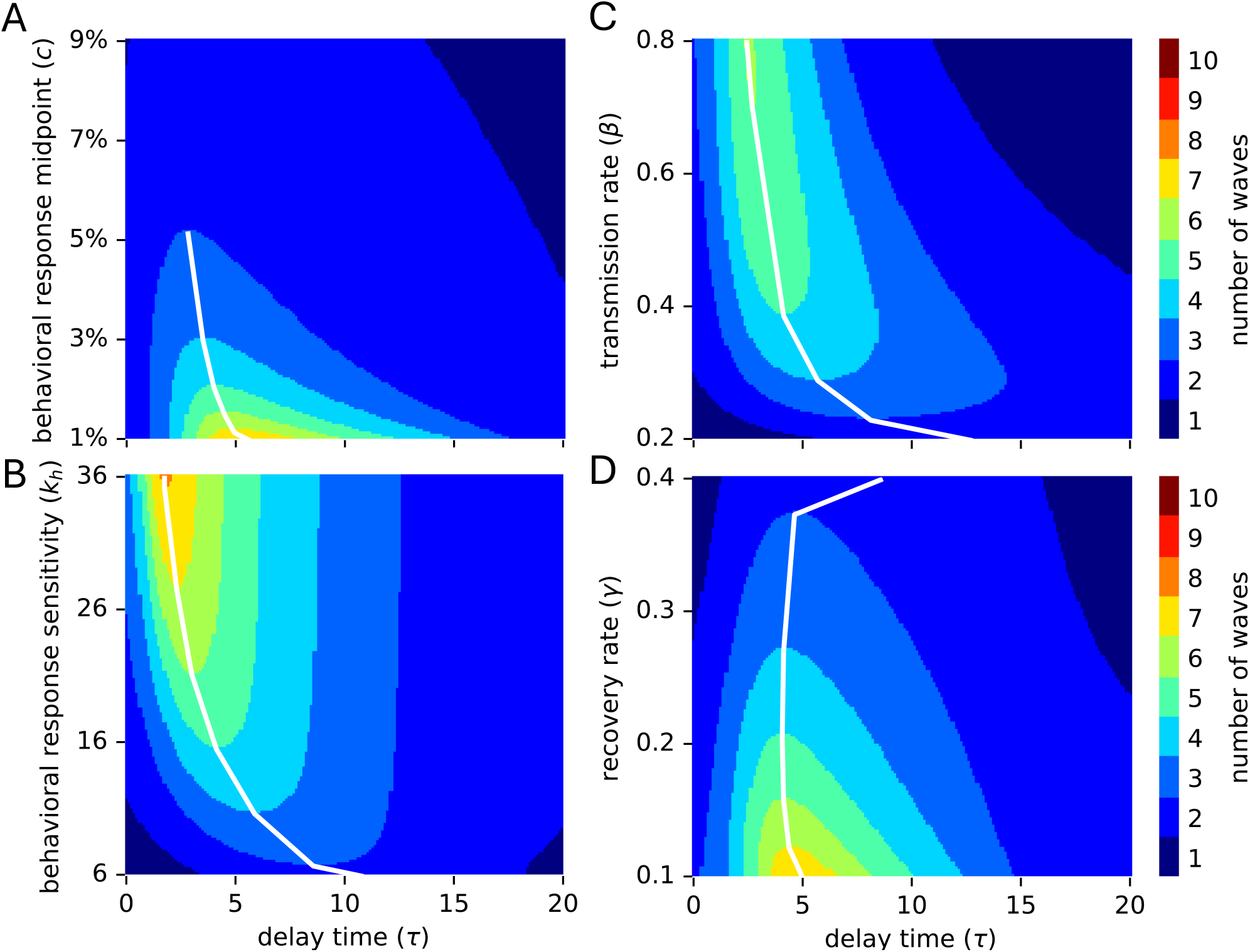
Two-dimensional sensitivity analysis, assuming SEIR-type dynamics with a latency period of 5 days. Exposed individuals are assumed to be non-infectious and not included in the prevalence, which determines the behavioral response. The number of epidemic waves is shown for a range of values for the delay parameter (*τ*, x-axis) and another model parameter (y-axis): (A) behavioral response midpoint *c*, (B) behavioral response sensitivity *k*_*h*_, (C) transmission rate *β*, (D) recovery rate *γ*. White lines connect the highest (in A,D) or lowest (in B,C) model parameter value and associated delay value that yields a specific number of multiple waves. All non-specified parameters are at their default values listed in Table 1.

**Figure S5.**
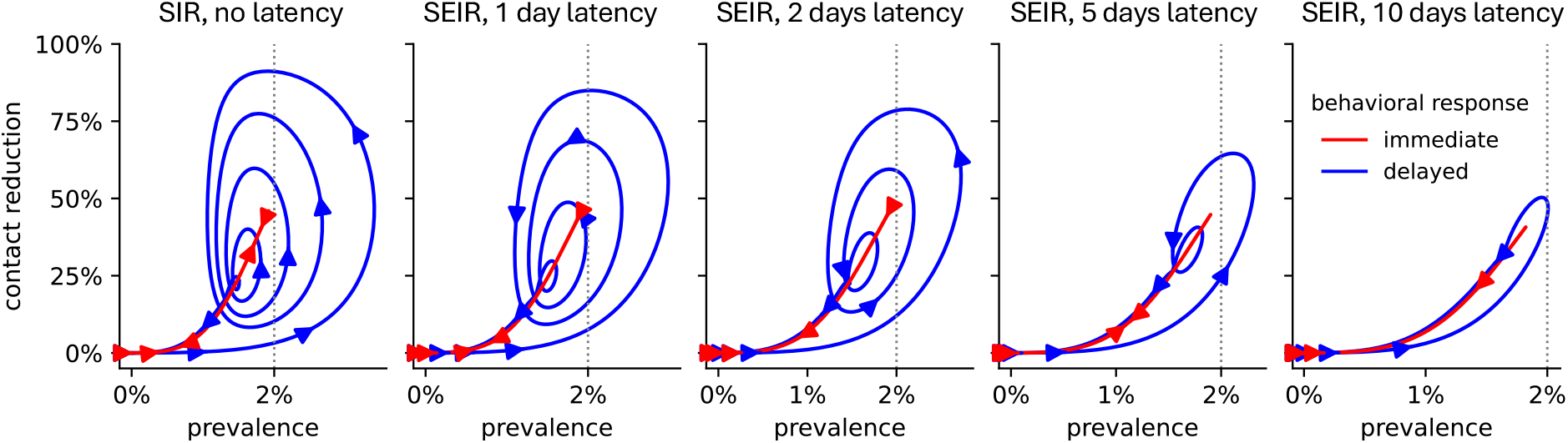
Effect of latency on the shape of disease dynamics. The trajectory of the prevalence and contact reduction under an immediate (red) and 5-day delayed (blue) behavioral response is shown. Exposed individuals are assumed to be non-infectious and not included in the prevalence, which determines the behavioral response. The arrows indicate the direction of the change over time. All non-specified parameters are at their default values listed in Table 1.

